# Test-retest repeatability of intravoxel incoherent motion (IVIM) measurements in the cervical cord

**DOI:** 10.1101/2024.04.06.24305341

**Authors:** Anna Lebret, Simon Lévy, Patrick Freund, Virginie Callot, Maryam Seif

## Abstract

This work aimed at assessing the reliability of intravoxel incoherent motion (IVIM) parameters sensitive to perfusion changes in the cervical cord by determining the test-retest variability across subjects and different post-processing fitting algorithms. IVIM test-retest scans were acquired in the cervical cord (C1-C3) of 10 healthy subjects on a 3T MRI scanner, with a 15-minute break in-between. IVIM parameters, including microvascular volume fraction (*F*), pseudo-diffusion coefficient (*D\**), blood flow-related coefficient (*F* · *D\**), and diffusion coefficient (*D*), were derived using voxel-wise and region of interest (ROI)-wise fits. The reliability of each IVIM parameter was determined with coefficients of variation (CV), intraclass correlation coefficients (ICC), Bland-Altman analysis and linear regression. To assess the effects of the different fitting approaches, a two-way repeated-measures analysis of variance (ANOVA) was conducted on the CVs calculated across fitting algorithms. Mean CVs of IVIM parameters calculated across subjects using the voxel-wise fit was lower in the white matter (WM) and grey matter (GM) (2.6% to 15.6%; 2.2% to 16.4%, respectively) compared with those calculated using the ROI-wise fit approach (WM: 4.5%-32.2%; GM: 3.4%-53.4%). The voxel-wise fit in the WM yielded higher ICC values (good-to-excellent, 0.71 – 0.97) compared to the ROI-wise fit approach (0.49 – 0.90). IVIM parameters, derived using the voxel-wise fitting approach, demonstrated a high reliability in the cervical cord. Robust IVIM metrics, observed across scans and subjects, can facilitate studies targeting perfusion impairment and pave the way to future clinical trials assessing perfusion impairment as a potential quantitative biomarker.

## Introduction

Intravoxel incoherent motion (IVIM) is a non-invasive quantitative MRI technique used to indirectly assess tissue micro-perfusion by determining the contributions of microcirculation and thermal diffusion to the MR signal decay^1^. It is based on the circulation of blood water molecules in the capillary bed as a pseudo-Brownian diffusion process, under the assumption of randomly oriented capillaries in the tissue^1^. IVIM MRI provides parameters indirectly sensitive to the microvascular volume (*F*), blood velocity (*D\**), blood flow (*F* · *D\**), and tissue diffusion (*D*)^2^. Previous reports have shown that IVIM is sensitive to changes in tissue perfusion^3–8^. This technique has thereby potential to study perfusion impairment and ensuing ischemia occurring in the damaged spinal cord in a variety of neurodegenerative diseases^9,10^. Characterization of perfusion dysfunction is crucial to better understand its role in the disease progression and severity. Previous studies have showed that different fitting approaches of the IVIM model^11,12^ can result into a large variability by applying different algorithms in organs of various perfusion levels^7,13–15^. The two most common fitting methods are the “two-step” or “segmented” approach and the “one-step” approach. These methods differ in the order of parameters estimation and have been previously explored in IVIM studies across various organs^11,13,16,17^. On the other hand, IVIM parameters in a specific region of interest (ROI) can be derived either using a voxel-wise fit approach (followed by averaging across the ROI) or a ROI-wise fit (fitting the average MR signal across the ROI). Thus, it is of great relevance to determine which fitting approach in IVIM modeling yields the most reliable outcomes in terms of robustness to noise and variability. While IVIM MRI has already been applied in different tissue types^3–8^, including the cervical cord^11^, its reliability is still understudied. Of note, cervical cord MRI measurements are limited by several challenges, including a low signal-to-noise ratio due to RF coil coverage compared to other organs such as the brain^18–20^, motion artifacts from both physiological and spinal cord-related movements^21^, and susceptibility artifacts induced by B_0_ inhomogeneities of different tissue types near spinal cord causing distortions and signal drop^18^. Determining IVIM parameters’ reliability in the cervical cord is therefore essential to quantify the minimum detectable pathophysiological changes and substantiate IVIM findings in future studies. The objective of this study was thus to determine the reliability of IVIM parameters calculated by the one-step and two-step fitting approach in the cervical cord, based on both voxel-wise and ROI-wise fitting methods.

## Methods

### Standard protocol approvals, registrations, and participant consents

The study protocol was designed according to the Declaration of Helsinki and approved by the local Ethics Committee (Kantonale Ethikkommission Zürich, EK-2018-00937). Informed written consent was obtained from each participant before study enrolment.

### Participants and MR acquisition

Ten healthy controls (mean age ± SD: 30.0 ± 5.4 years, 4 females) were scanned twice, with a 15-minute break out of the scanner between the sessions. The exclusion criteria for the study enrollment were prior operation to the cervical spine, MRI contraindication, pre-existing neurological condition, and age under 18 or above 75 years old. MRI data were acquired on a 3T MRI scanner (MAGNETOM Prisma, Siemens Healthineers, Erlangen, Germany) with a Siemens Healthineers 64-channel head and neck radio-frequency coil. A stifneck collar (Laerdal Medicals, Norway) was used on every subject to minimize motion artifacts in the inferior-superior direction.

### Image acquisition protocol

Study participants underwent a protocol composed of a sagittal T2-weighted turbo spin echo (TSE) sequence to localize the cervical levels, an axial T2*-weighted 3D multi-echo gradient-echo sequence^22^ to segment grey and white matter, and a vendor product cardiac-gated IVIM axial 2D-RF diffusion-weighted spin-echo EPI ZOOMit (Zonally-magnified Oblique Multislice) sequence with a trigger delay of 100 ms (using the Siemens standard pulse oximeter), a 34×108 matrix size, 9 slices and 3 concatenations (3 slices/cardiac cycle), 14 b-values ranging from 0 to 650 s/mm^2^ with an increment of 50 s/mm^2^ ([0; 50; 100; 150; 200; 250; 300; 350; 400; 450; 500; 550; 600; 650])^23^ with 20 repetitions per b-value in three in-plane diffusion encoding directions to assess perfusion (60deg, 180deg, -60deg). Diffusion-encoding directions were restricted to the axial plane, as the estimation of IVIM parameters along the inferior-superior spinal axis has been shown previously to be compromised by the high water diffusivity coefficient in that direction^23^. The IVIM acquisitions were split into forward and reverse phase encoding directions for subsequent distortion correction (10 repetitions/b-value in each phase-encoding direction). The actual acquisition time depended on the subject’s heartbeat and varied between 6 and 10 minutes for each phase-encoding direction. All scans were acquired in the cervical cord covering C1-C3 levels. The total nominal acquisition time per scan-rescan session was ca. 39 minutes (depending on the heart rate). MRI parameters of the different sequences are reported in Table 1.

**Table 1:**
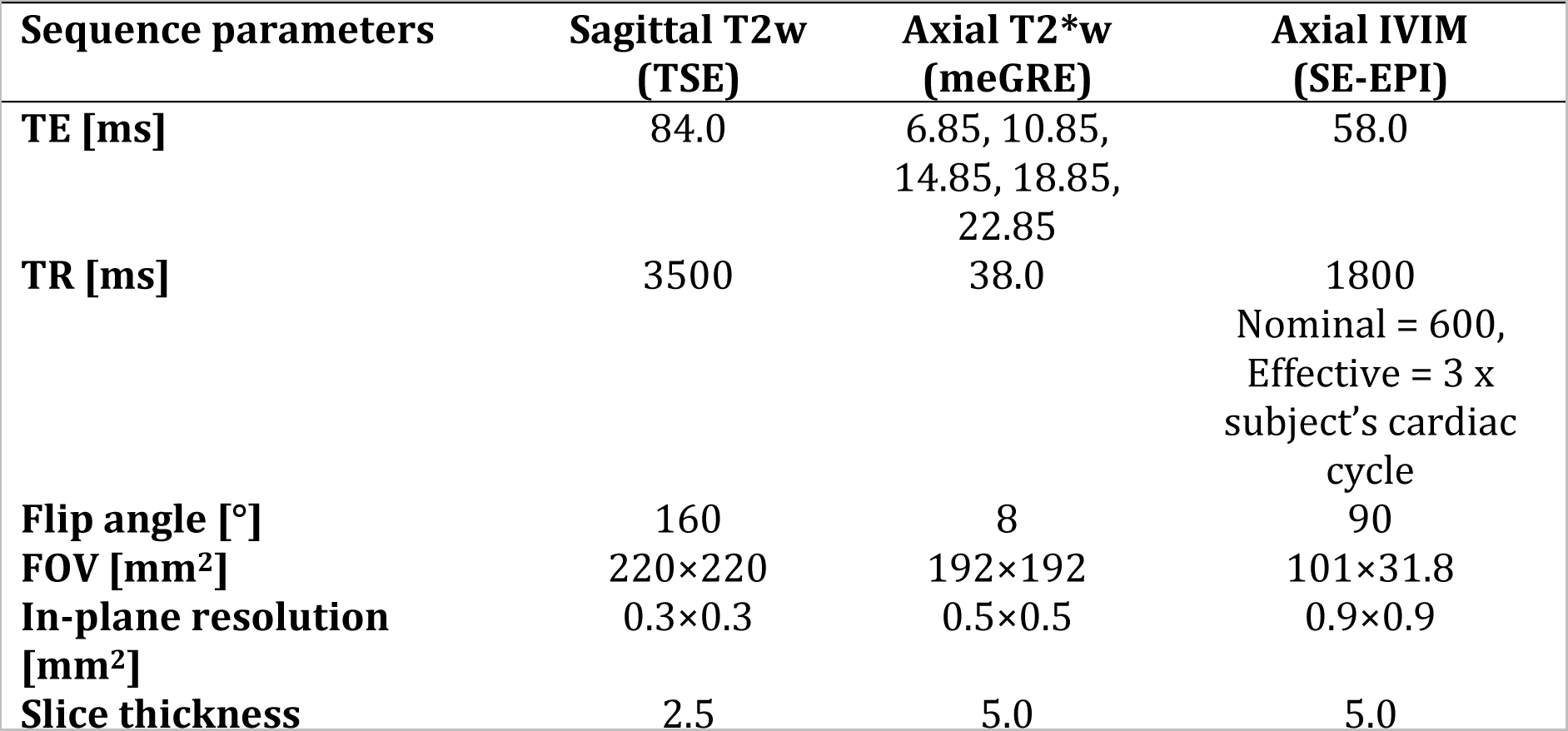
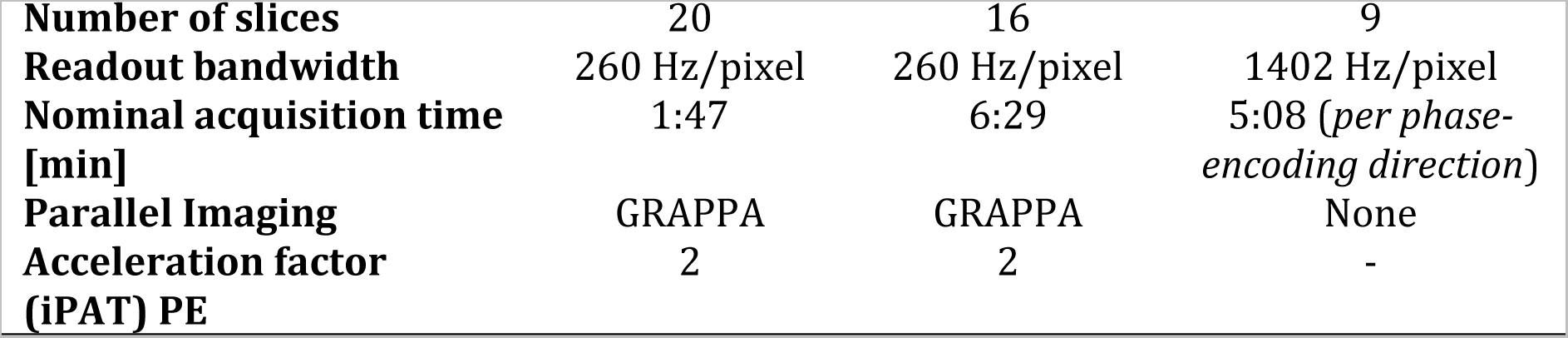
MRI scan parameters of the protocol sequences. GRAPPA: generalized autocalibration partially parallel acquisition.

| Sequence parameters | Sagittal T2w (TSE) | Axial T2*w (meGRE) | Axial IVIM (SE-EPI) |
| --- | --- | --- | --- |
| TE [ms] | 84.0 | 6.85, 10.85, 14.85, 18.85, 22.85 | 58.0 |
| TR [ms] | 3500 | 38.0 | 1800<br>Nominal = 600,<br>Effective = 3 x<br>subject's cardiac<br>cycle |
| Flip angle [°] | 160 | 8 | 90 |
| FOV [mm<sup>2</sup>] | 220×220 | 192×192 | 101×31.8 |
| In-plane resolution [mm<sup>2</sup>] | 0.3×0.3 | 0.5×0.5 | 0.9×0.9 |
| Slice thickness | 2.5 | 5.0 | 5.0 |
| **Number of slices** | 20 | 16 | 9 |
| **Readout bandwidth** | 260 Hz/pixel | 260 Hz/pixel | 1402 Hz/pixel |
| **Nominal acquisition time [min]** | 1:47 | 6:29 | 5:08 (*per phase-encoding direction*) |
| **Parallel Imaging** | GRAPPA | GRAPPA | None |
| **Acceleration factor (iPAT) PE** | 2 | 2 | - |

### Image processing

IVIM images were processed using the IVIM toolbox^11^. The processing steps consisted of denoising of each phase-encoding direction using a local principal component analysis algorithm^24^, removal of Gibbs artefacts using the local sub-voxel-shifts method^25,26^ (*DIPY*^27^), and finally motion (*sct_dmri_moco*^28^) and EPI-related distortion correction (*FSL topup*^29^).

Prior to fitting the IVIM signal, voxel-wise SNR maps were determined to evaluate data quality and compare SNR values to previous investigations assessing required SNR for model accuracy^11^. The maps were generated based on the distortion-corrected images acquired at b_0_ and calculated as the ratio of the mean signal across repetitions to the standard deviation across repetitions, in a voxel-wise manner^11,30^. To represent the signal fed to the fitting algorithm, the SNR values were multiplied by a factor √N_rep_(representing the number of repetitions). SNR values were extracted in subject space within the eroded spinal cord mask (to minimize partial volume effect with the CSF) and averaged across C1-C3 levels and subjects for each scanning session.

### IVIM signal fitting

The IVIM signal is commonly described by the following biexponential decay model^1^:

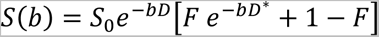

where *F* is the microvascular volume fraction, *D\** is the pseudo-diffusion coefficient (proportional to blood velocity in the capillaries^2^), and *D* is the diffusion coefficient. Furthermore, *F* · *D\** can be proportionally related to blood flow^2^.

To assess the robustness of the fit and the reliability of IVIM measurements obtained with different fitting approaches, IVIM maps were calculated with a voxel-wise and ROI-wise fit, using two different standard algorithms, namely a constrained one-step method and a constrained two-step “segmented” method^11^.

### One-step and two-step algorithms

Briefly, in the one-step algorithm^31^, a prior estimation of *D* was performed for boundaries determination using only a mono-exponential fit based on b-values above 400 [s/mm^2^] and using the *Conjugate Gradient* optimization method. The fit of all parameters was then performed based on all b-values (0 to 650 [s/mm^2^]) using the *Differential Evolution* optimization method^32^ (to escape local optima) and constraining *D* between 1.5×10^-4^ [mm^2^/s] and 54×10^-4^ [mm^2^/s]. The other parameters were confined in a range of values published in the literature for the brain: *F* [%]: [0; 20]; *D\** [mm^2^/s ×10^-3^]: [0.3; 50]. In a final step, fine-tuning of the parameters with tighter boundaries (constraining the parameter estimation to 95% - 105% of the previously estimated values) was conducted.

In the two-step method, the IVIM parameters are estimated in two different steps. *D* was first estimated using a mono-exponential fit based on b-values above 400 [s/mm^2^]. The fit of *F* and *D\** was then performed on all b-values as described for the one-step method, keeping *D* fixed to the value estimated in the first step. The last step consisted of fine-tuning of *F* and *D\**, constraining the value estimation with tighter boundaries (95% - 105% of the estimation obtained at the previous step), while *D* remained fixed to its initial estimation.

### Voxel-wise and ROI-wise fits

For the voxel-wise approach, the IVIM signal was fitted (using the one-step and two-step algorithms) in each voxel of the spinal cord segmentation (produced with *sct_deepseg_sct*^33^), resulting in voxel-wise IVIM maps in each diffusion-encoding direction.

White matter and grey matter were investigated separately in the ROI-wise approach. Probabilistic masks were calculated from the white matter atlas registered to the diffusion images (generated during the registration step, see next section) and eroded at the periphery of the spinal cord to avoid partial volume effect with the cerebrospinal fluid (CSF). The signal value in each ROI was obtained by calculating the weighted average of the diffusion signal values in each voxel with the probability value of the probabilistic mask in that voxel, per slice and per b-value. This ROI-wise signal decay was then fitted (using either the one-step or two-step algorithms) resulting in ROI-wise IVIM values per slice in each diffusion-encoding direction.

### Registration to template

The white and grey matter probabilistic masks were needed to generate an average signal value per slice in the ROI-wise approach and extract the IVIM metrics. Thus, the IVIM images and maps were registered to the PAM50 template^34^ and white matter atlas^35^ with an intermediate registration step performed on T2*-weighted images to account for the subject’s variability in the grey matter shape (Figure 1, step 3). The IVIM maps in the 60° and -60° diffusion-encoding directions were registered to the 180° one and then averaged across directions (Figure 1, step 1C). Finally, values of the IVIM parameters were extracted from the individual maps (averaged across directions), in subject space, averaged across C1-C3 vertebral segments, in the ROIs, for both the ROI-wise and voxel-wise approaches (Figure 1, step 4). The regions of interest were eroded before metric extraction at the cord periphery, with *fslmaths* from the FSL library (v6.0.5, Analysis Group, FMRIB, Oxford, UK)^36^, using a sphere kernel of radius 1, to minimize partial volume effect with the CSF. An overview of the processing pipeline is shown in Figure 1.

**Figure 1:**
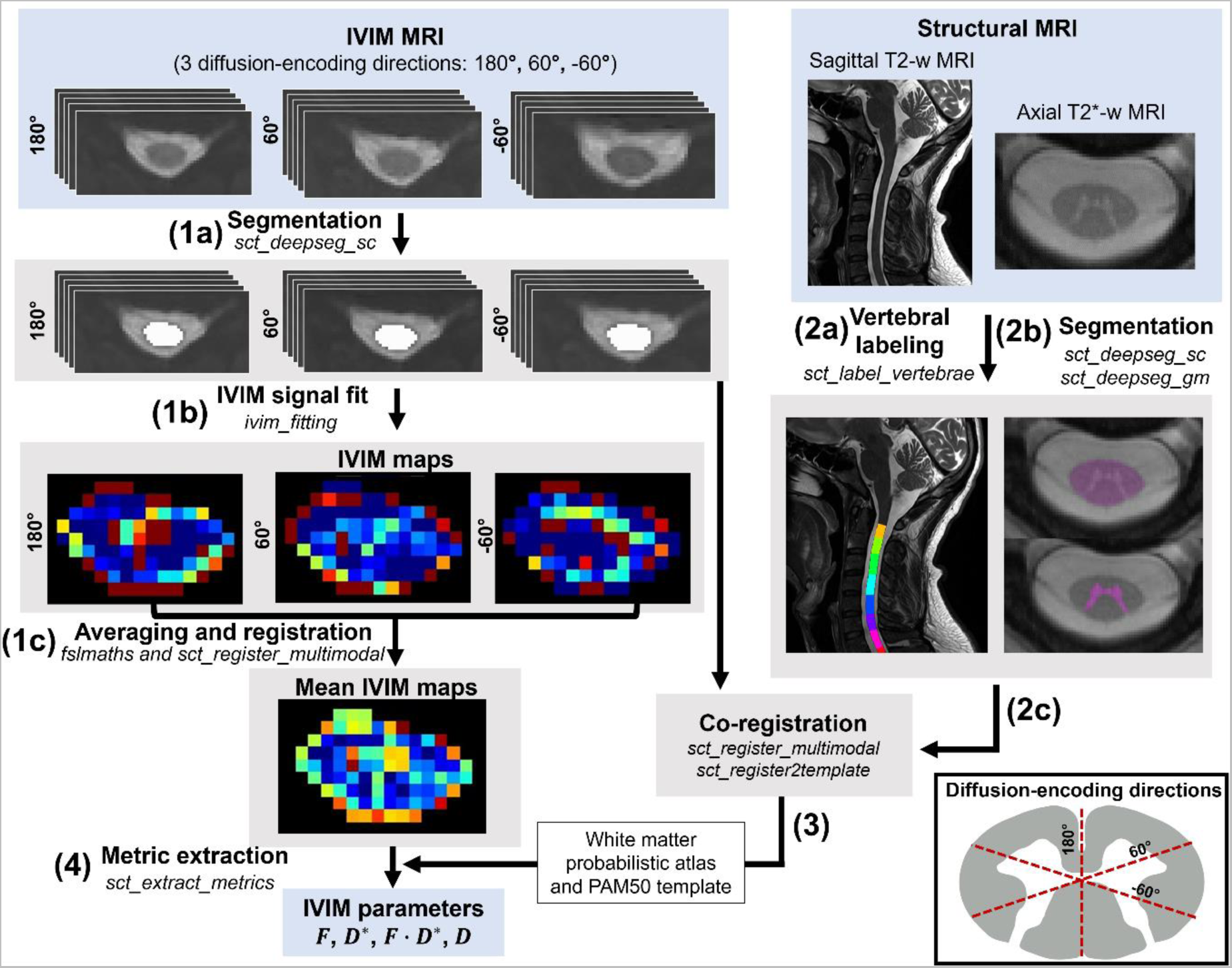
IVIM post-processing pipeline using the IVIM toolbox^11^. IVIM MRI: (1a) segmentation of the spinal cord is conducted on each individual diffusion-encoding direction (i.e., 180°, 60°, -60°). (1b) IVIM maps (here *F* maps are shown for illustration) are generated from the distortion-corrected diffusion images using the voxels provided by the cord segmentations in each of the three diffusion-encoding directions and then (1c) averaged across directions (after registration of the IVIM images to the 180° diffusion-encoding direction). Structural MRI: (2a) vertebral labeling on sagittal T2-weighted MRI and (2b) segmented spinal cord and grey matter on the axial T2*-weighted images are obtained and (2c) further used for the co-registering to the IVIM images. As a final step, (4) IVIM parameters are extracted in each slice in the white and grey matter after (3) registration of the maps to the PAM50 template and its white matter atlas, using warping fields derived from the co-registration of the IVIM images to the structural images (at step 2c). Note that the averaging across repetitions per b-value and the distortion correction are not illustrated here and occurred before the registration pipeline. The averaging across slices corresponding to C1-C3 levels (from which the test-retest differences, CV and ICC were then derived) are not illustrated either. Visualization of the diffusion-encoding directions is provided in the box in the bottom right corner.

### Test-retest statistical analysis

The test-retest repeatability was determined for the four fitting approaches investigated (voxel-wise one-step, voxel-wise two-step, ROI-wise one-step, and ROI-wise two-step), using the following standard reliability measures:

The within-subject (ws) coefficients of variation (CV), estimating the variability across two scans for each subject, were calculated by the standard deviation (*σ_ws_*) over the mean (*µ_ws_*) across both scanning sessions for each subject:

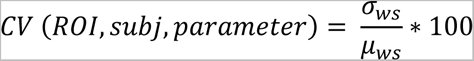

where *ROI* represents the different regions of interest, *subj* is the subject participating to the study, and *parameter* is each parameter of the IVIM model (*F*, *D\**, *F* · *D\**, and *D*). To assess the effect of different fitting approaches (voxel-wise vs. ROI-wise), and fitting algorithms (one-step vs. two-step) on the CVs, a two-way ANOVA was performed. The dependent variable was the CVs values, while the independent variables were the different fitting approaches and the fitting algorithms as the two categorical variables. The analysis was within each ROI with a significance level set at α = 0.05.

The intraclass correlation coefficient (ICC) was applied to determine the test-retest reliability as a ratio of true variance (between the subjects) over the true variance plus the variance due to measurement errors^37^. The ICC implementation was based on MATLAB (version 9.13.0 (R2022b), the MathWorks, Inc., case 3A, absolute agreement)^38^ and the definition of McGraw and Wong^39^ for each parameter in each ROI. For the interpretation of the ICC values, we used the common scale introduced by Cicchetti^40^, Cicchetti and Sparrow^41^, and Fleiss^42^: *poor < 0.4 < fair < 0.6 < good < 0.75 < excellent*.

Next, a Bland-Altman analysis was conducted to define the mean difference between the sessions (i.e., bias) against the average value across sessions, along with the 95% limits of agreements (average difference ± 1.96 standard deviation of the mean difference).

Finally, Pearson linear correlations were performed between values obtained in the first and second sessions. A better test-retest repeatability is therefore represented by lower CVs, higher ICC, lower bias, and narrower limits of agreements in Bland-Altman analysis. Statistical analysis was performed using R software (version 4.1.2, R Core Team^43^), aside from the ICC computation done in MATLAB.

## Results

### SNR values in individual diffusion-encoding directions

Table 2 presents the mean, standard deviation, minimum, and maximum SNR values averaged across C1-C3 levels and subjects on the lowest b-value images on the distortion-corrected images (input of the fitting algorithm). The values are reported for each of the three diffusion-encoding directions and each scanning session. SNR values remained similar across the various diffusion-encoding directions and scanning sessions.

**Table 2:**
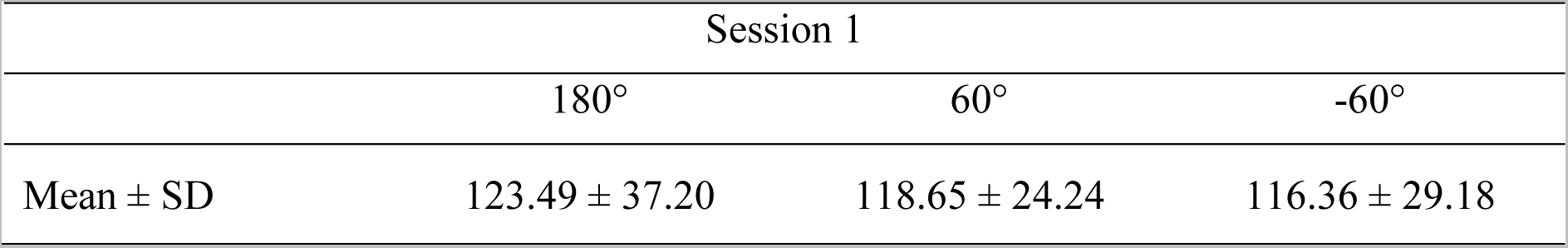
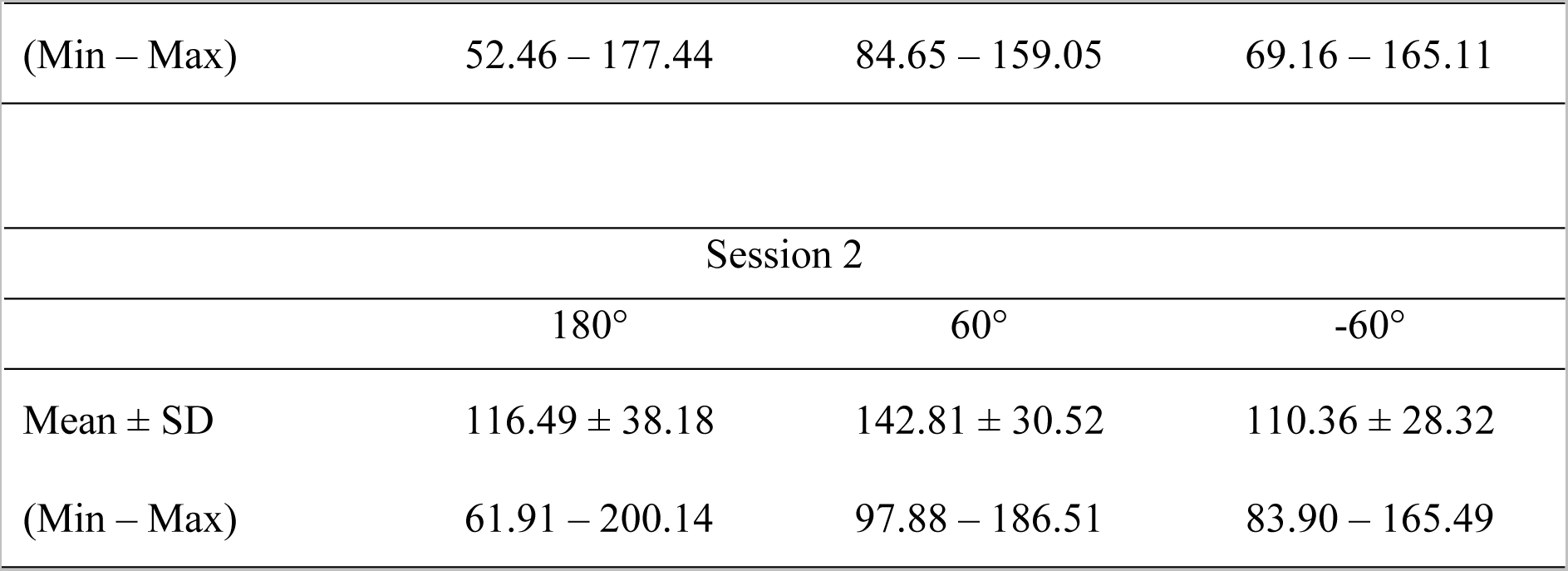
Mean and standard deviation of SNR values at b_0_ of the distortion-corrected IVIM images within the eroded spinal cord mask, averaged across C1-C3 levels and subjects, and value ranges, for each diffusion-encoding direction (180°, 60°, and -60°) and scanning session separately.

### Mean IVIM values in the cervical cord

Figure 2 shows IVIM maps (*F*, *D\**, *F* · *D\**, and *D*) calculated in the voxel-wise fit using the one-step algorithm, averaged across subjects and C1-C3 levels. The microvascular volume fraction *F* appeared distinctly higher in the anterior and intermediate parts of the grey matter and highlights the greater vascularization of this structure of the spinal cord. The mean and standard deviation maps across subjects showed high similarity between the two sessions compared to the single subject maps, indicating a qualitatively good repeatability of the IVIM parameters averaged at the group level. The mean values of each IVIM parameter, averaged across C1-C3 levels, in the white and grey matter calculated with the voxel-wise and ROI-wise fits in each session are reported in Table 3.

**Figure 2:**
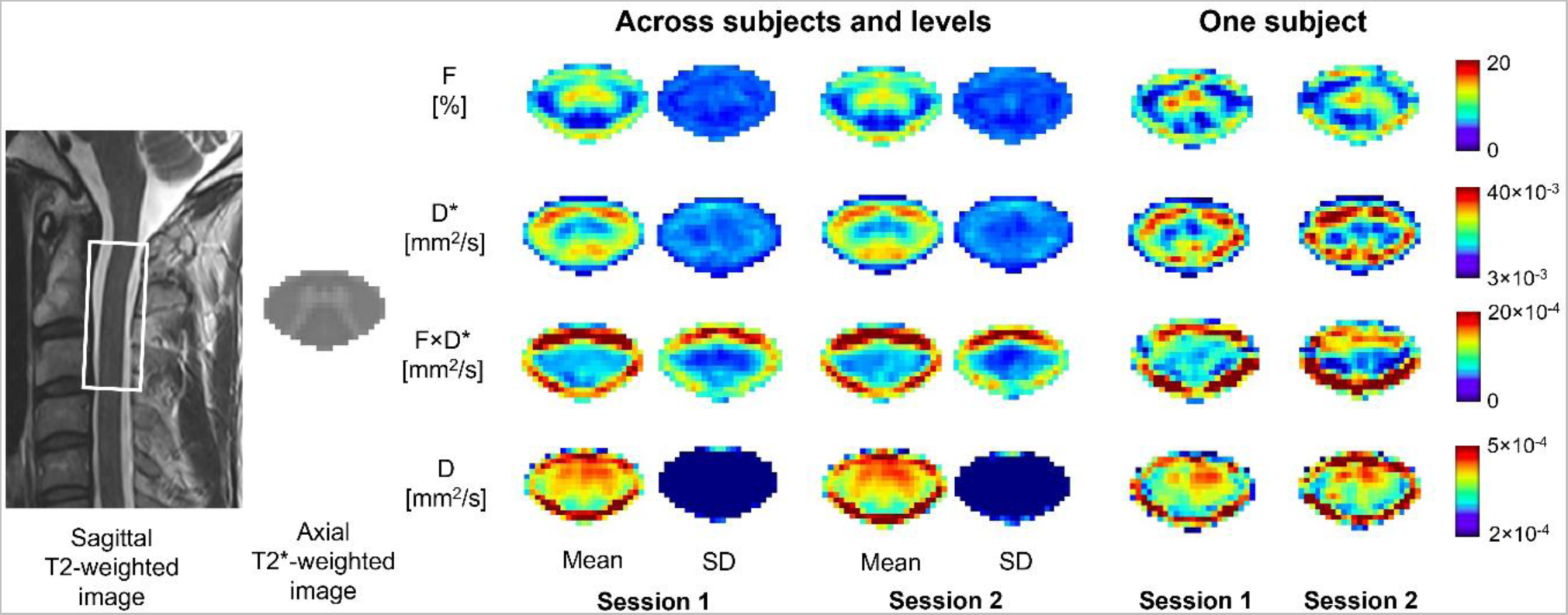
Voxel-wise one-step fit mean and standard deviation (SD) maps of the IVIM parameters (microvascular fraction (*F*), blood velocity-related coefficient (*D\**), blood flow-related coefficient (*F* · *D\**), and diffusion coefficient (*D*)) in the template space, shown across subjects and for one subject, for both scanning sessions. Maps are averaged across C1-C3 vertebral levels and diffusion-encoding directions.

**Table 3:** Mean and standard deviation values of IVIM parameters across participants. Values of IVIM parameters obtained in the white matter (WM) and grey matter (GM) for both sessions and mean (averaged across subjects) of the relative differences between the average values of the two sessions, using the one-step and two-step algorithms with (A) the voxel-wise fit and (B) the ROI-wise fit. S1: Session 1; S2: Session 2; Δ: average (across subjects) of the relative differences (session 1 – session 2) in percentage of session 1.

| | $F$ [%] | | | $D^*$ [$\text{mm}^2/\text{s} \times 10^{-3}$] | | | $F \cdot D^*$ [$\text{mm}^2/\text{s} \times 10^{-4}$] | | | $D$ [$\text{mm}^2/\text{s} \times 10^{-4}$] | | |
| --- | --- | --- | --- | --- | --- | --- | --- | --- | --- | --- | --- | --- |
| | S 1 | S 2 | $\Delta$ [%] | S 1 | S2 | $\Delta$ [%] | S 1 | S 2 | $\Delta$ [%] | S 1 | S 2 | $\Delta$ [%] |
| *one-step* |  |  |  |  |  |  |  |  |  |  |  |  |
| WM | 5.45±0.75 | 5.49±0.94 | 11.18 | 24.41±2.61 | 24.32±2.89 | 4.39 | 10.16±2.67 | 10.32±3.52 | 11.66 | 3.74±0.49 | 3.77±0.55 | 3.73 |
| GM | 9.61±1.55 | 9.71±1.39 | 16.98 | 17.14±2.89 | 16.82±3.45 | 12.77 | 8.48±2.08 | 8.63±2.17 | 12.12 | 4.17±0.42 | 4.17±0.48 | 3.08 |
| | $F$ [%] | | | $D^*$ [$\text{mm}^2/\text{s} \times 10^{-3}$] | | | $F \cdot D^*$ [$\text{mm}^2/\text{s} \times 10^{-4}$] | | | $D$ [$\text{mm}^2/\text{s} \times 10^{-4}$] | | |
| | S 1 | S 2 | $\Delta$ [%] | S 1 | S2 | $\Delta$ [%] | S 1 | S 2 | $\Delta$ [%] | S 1 | S 2 | $\Delta$ [%] |
| *two-step* |  |  |  |  |  |  |  |  |  |  |  |  |
| WM | 4.03±1.05 | 4.18±1.29 | 23.67 | 24.38±3.64 | 24.07±3.20 | 8.57 | 7.50±2.28 | 7.62±2.85 | 11.28 | 4.23±0.46 | 4.25±0.49 | 5.74 |
| GM | 6.60±1.40 | 6.80±1.53 | 21.93 | 15.47±3.94 | 15.54±3.54 | 24.24 | 6.74±1.88 | 6.78±1.76 | 13.94 | 4.62±0.40 | 4.62±0.45 | 4.68 |

(B) ROI-wise
| | $F$ [%] | | | $D^*$ [$\text{mm}^2/\text{s} \times 10^{-3}$] | | | $F \cdot D^*$ [$\text{mm}^2/\text{s} \times 10^{-4}$] | | | $D$ [$\text{mm}^2/\text{s} \times 10^{-4}$] | | |
| --- | --- | --- | --- | --- | --- | --- | --- | --- | --- | --- | --- | --- |
| | S 1 | S 2 | $\Delta$ [%] | S 1 | S2 | $\Delta$ [%] | S 1 | S 2 | $\Delta$ [%] | S 1 | S 2 | $\Delta$ [%] |
| *one-step* |  |  |  |  |  |  |  |  |  |  |  |  |
| WM | 2.66±1.09 | 2.76±1.95 | 46.99 | 29.21±10.39 | 29.57±8.18 | 26.77 | 4.29±2.49 | 5.02±2.56 | 37.77 | 3.82±0.48 | 3.88±0.57 | 6.50 |
| GM | 11.45±4.82 | 11.03±3.25 | 40.05 | 11.68±9.19 | 13.56±8.39 | 100.48 | 4.62±2.11 | 4.64±1.54 | 26.27 | 3.98±0.41 | 4.04±0.34 | 8.99 |
| | $F$ [%] | | | $D^*$ [$\text{mm}^2/\text{s} \times 10^{-3}$] | | | $F \cdot D^*$ [$\text{mm}^2/\text{s} \times 10^{-4}$] | | | $D$ [$\text{mm}^2/\text{s} \times 10^{-4}$] | | |
| | S 1 | S 2 | $\Delta$ [%] | S 1 | S2 | $\Delta$ [%] | S 1 | S 2 | $\Delta$ [%] | S 1 | S 2 | $\Delta$ [%] |
| *two-step* |  |  |  |  |  |  |  |  |  |  |  |  |
| WM | 1.73±0.91 | 2.16±1.27 | 57.07 | 24.20±12.48 | 21.23±8.96 | 38.16 | 2.63±1.53 | 2.91±2.28 | 43.24 | 4.00±0.46 | 4.05±0.48 | 7.10 |
| GM | 5.08±1.21 | 5.29±1.66 | 27.80 | 9.07±7.41 | 10.55±7.91 | 120.76 | 3.48±2.28 | 3.10±1.03 | 44.53 | 4.60±0.42 | 4.59±0.42 | 4.83 |

### Test-retest repeatability of IVIM parameters

#### Comparing voxel-wise with ROI-wise fitting approaches

The within-subject CVs and the ICC calculated applying the voxel-wise and ROI-wise fits are reported in Table 4. The two-way ANOVA analysis resulted in a statistically significant difference across the mean of CVs across the different fitting approaches (voxel-wise vs. ROI-wise), where CVs calculated from the voxel-wise fitting approach were significantly lower compared to the ROI-wise fit (WM: *F*: p = 0.001, *D\**: p = 5.3e-5, *F* · *D\**: p = 0.005; GM: *D\**: 0.0003, *F* · *D\**: p = 0.002, *D*: p = 0.01), except for *D* in the white matter (p = 0.1) and for *F* in the grey matter (p = 0.07).

**Table 4:**
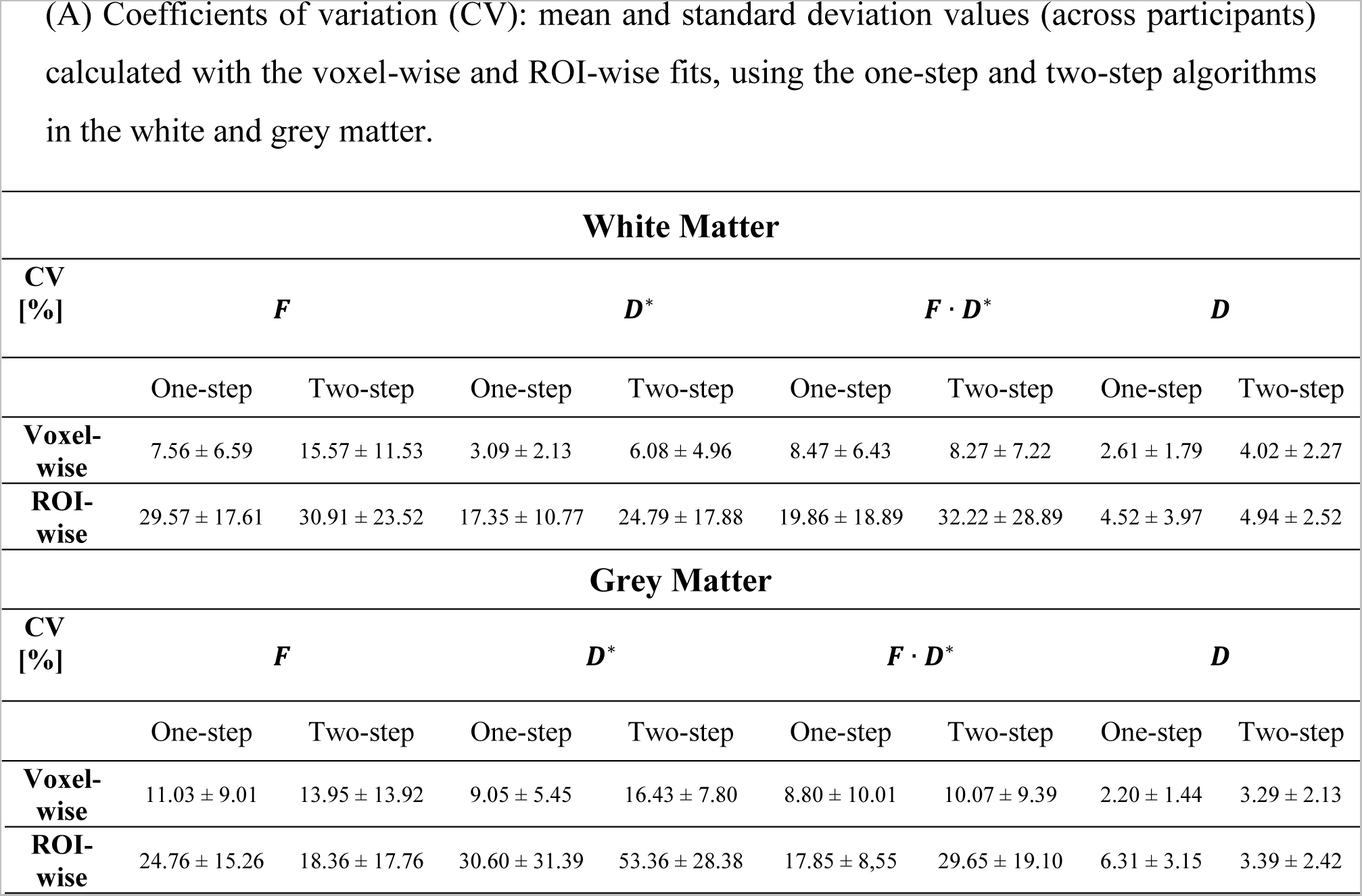
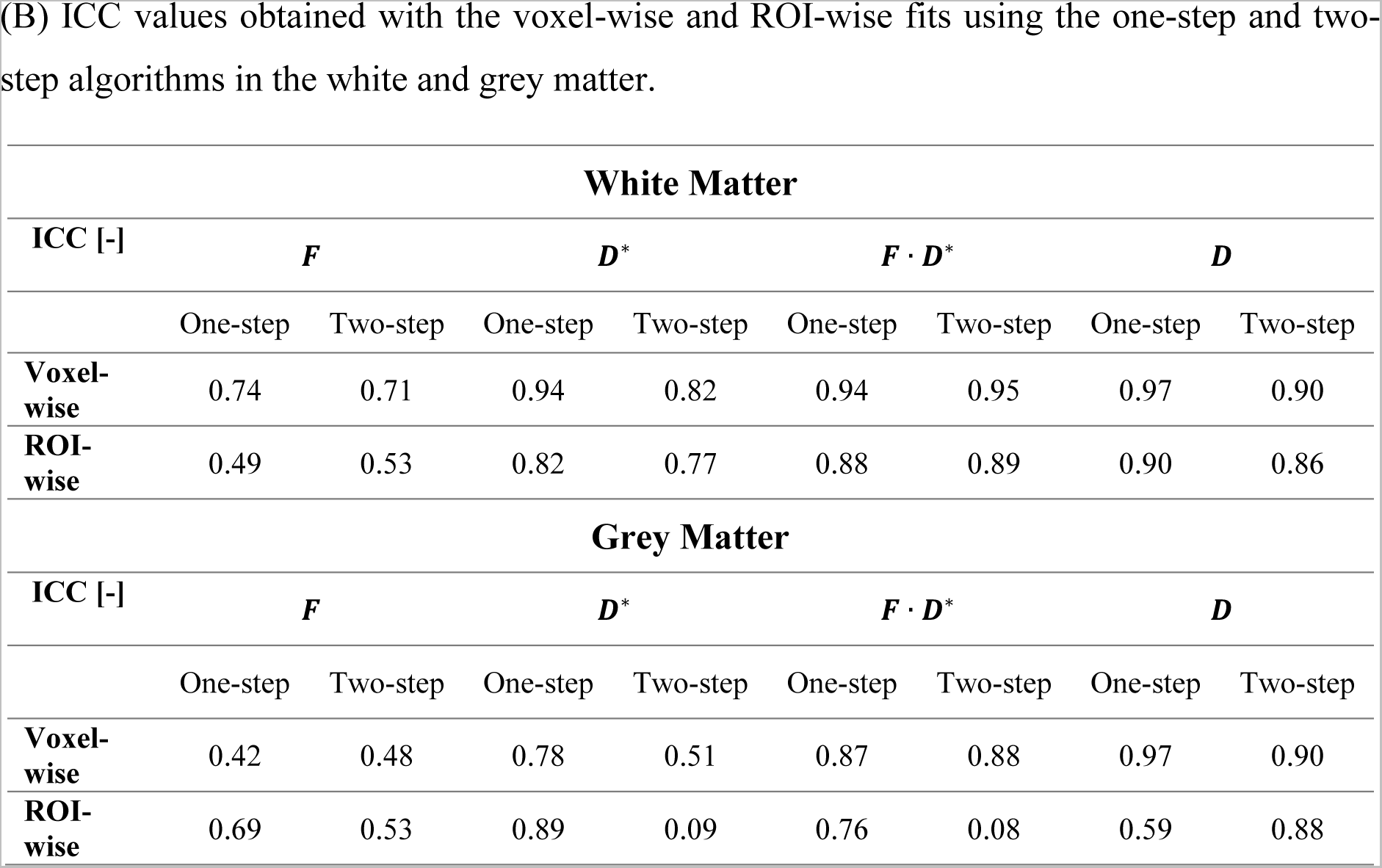
Test-retest repeatability of IVIM parameters. Values of (A) within-subject coefficients of variation (CV) averaged across subjects, and (B) ICC for the different IVIM parameters with the voxel-wise and ROI-wise fits using the one-step and two-step algorithms, in the white matter (WM) and grey matter (GM).

| White Matter |  |  |  |  |  |  |  |  |
| --- | --- | --- | --- | --- | --- | --- | --- | --- |
| CV<br>[%] | $F$ | | $D^*$ | | $F \cdot D^*$ | | $D$ | |
|  | One-step | Two-step | One-step | Two-step | One-step | Two-step | One-step | Two-step |
| Voxel-wise | $7.56 \pm 6.59$ | $15.57 \pm 11.53$ | $3.09 \pm 2.13$ | $6.08 \pm 4.96$ | $8.47 \pm 6.43$ | $8.27 \pm 7.22$ | $2.61 \pm 1.79$ | $4.02 \pm 2.27$ |
| ROI-wise | $29.57 \pm 17.61$ | $30.91 \pm 23.52$ | $17.35 \pm 10.77$ | $24.79 \pm 17.88$ | $19.86 \pm 18.89$ | $32.22 \pm 28.89$ | $4.52 \pm 3.97$ | $4.94 \pm 2.52$ |
| Grey Matter |  |  |  |  |  |  |  |  |
| CV<br>[%] | $F$ | | $D^*$ | | $F \cdot D^*$ | | $D$ | |
|  | One-step | Two-step | One-step | Two-step | One-step | Two-step | One-step | Two-step |
| Voxel-wise | $11.03 \pm 9.01$ | $13.95 \pm 13.92$ | $9.05 \pm 5.45$ | $16.43 \pm 7.80$ | $8.80 \pm 10.01$ | $10.07 \pm 9.39$ | $2.20 \pm 1.44$ | $3.29 \pm 2.13$ |
| ROI-wise | $24.76 \pm 15.26$ | $18.36 \pm 17.76$ | $30.60 \pm 31.39$ | $53.36 \pm 28.38$ | $17.85 \pm 8.55$ | $29.65 \pm 19.10$ | $6.31 \pm 3.15$ | $3.39 \pm 2.42$ |
| ICC [-] | $F$ | | $D^*$ | | $F \cdot D^*$ | | $D$ | |
|  | One-step | Two-step | One-step | Two-step | One-step | Two-step | One-step | Two-step |
| **Voxel-wise** | 0.74 | 0.71 | 0.94 | 0.82 | 0.94 | 0.95 | 0.97 | 0.90 |
| **ROI-wise** | 0.49 | 0.53 | 0.82 | 0.77 | 0.88 | 0.89 | 0.90 | 0.86 |

| Grey Matter |  |  |  |  |  |  |  |  |
| --- | --- | --- | --- | --- | --- | --- | --- | --- |
| ICC [-] | $F$ | | $D^*$ | | $F \cdot D^*$ | | $D$ | |
|  | One-step | Two-step | One-step | Two-step | One-step | Two-step | One-step | Two-step |
| **Voxel-wise** | 0.42 | 0.48 | 0.78 | 0.51 | 0.87 | 0.88 | 0.97 | 0.90 |
| **ROI-wise** | 0.69 | 0.53 | 0.89 | 0.09 | 0.76 | 0.08 | 0.59 | 0.88 |

#### Comparing one-step with two-step fitting algorithms

While the one-step fitting approach resulted in lower CVs compared to the two-step approach, this difference was not statistically significant. The diffusion coefficient (*D*) showed the lowest CVs in all fitting scenarios (Figure 3). The ICC values (Figure 4) obtained in the white matter were higher with the voxel-wise one-step fitting approach (ranged from 0.74 to 0.97) compared to all other tested scenarios, except for the *F* · *D\** parameter (ICC: 0.95 vs. 0.94 with the voxel-wise one-step approach). In the grey matter, higher ICC values were obtained for *F* and *D\** using the ROI-wise fit, whereas voxel-wise yielded better ICC for *F* · *D\** and *D*.

**Figure 3:**
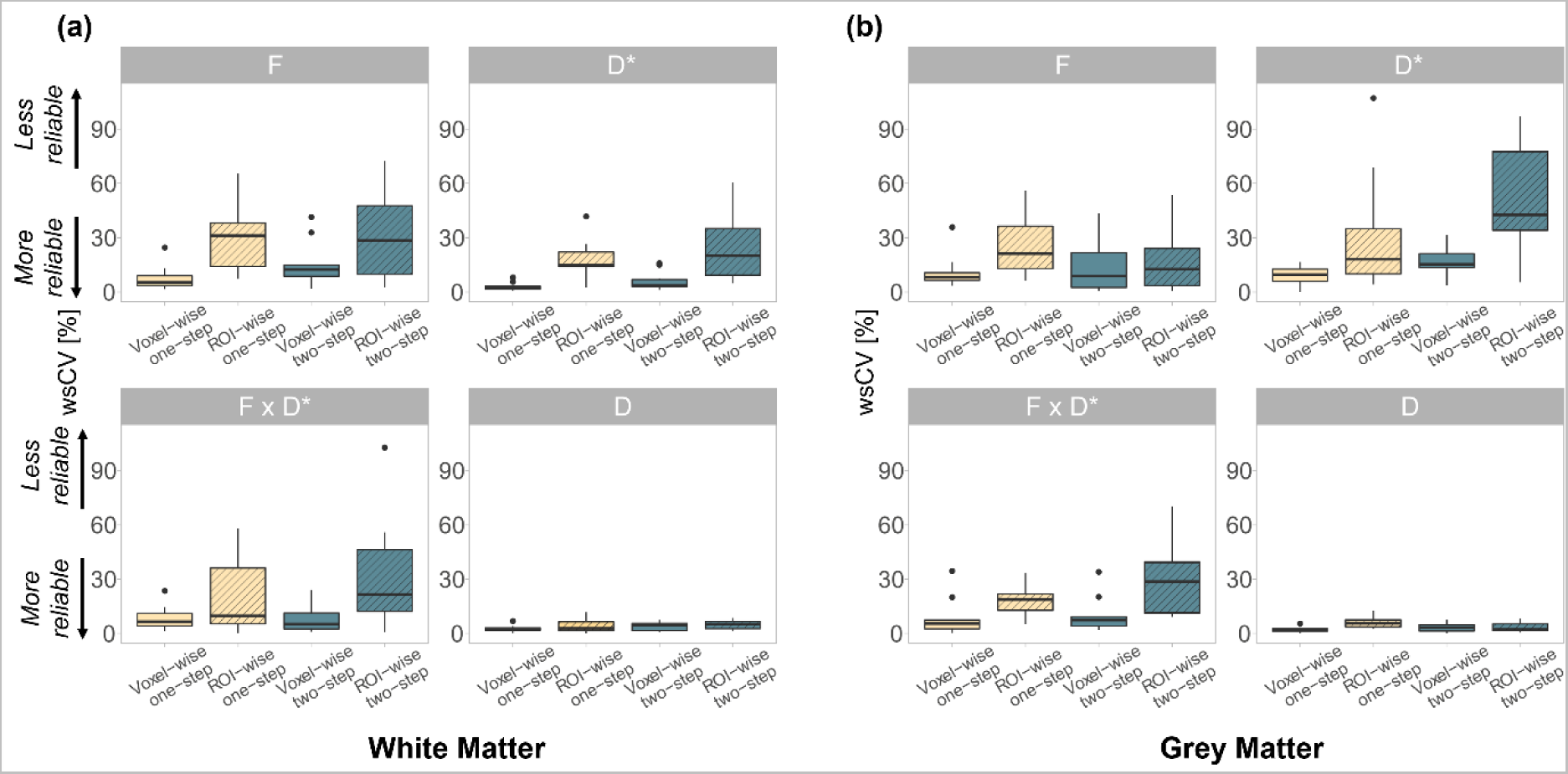
Within-subject coefficients of variation (wsCV) of the IVIM parameters obtained with the voxel-wise and ROI-wise fits, using the one-step and two-step algorithms in the (a) white matter and (b) grey matter. The box extends from the 1^st^ to the 3^rd^ quartiles, whiskers extend to the smallest and largest value no further than 1.5 times the inter-quartile range, while dots represent outlying values beyond the end of the whiskers.

**Figure 4:**
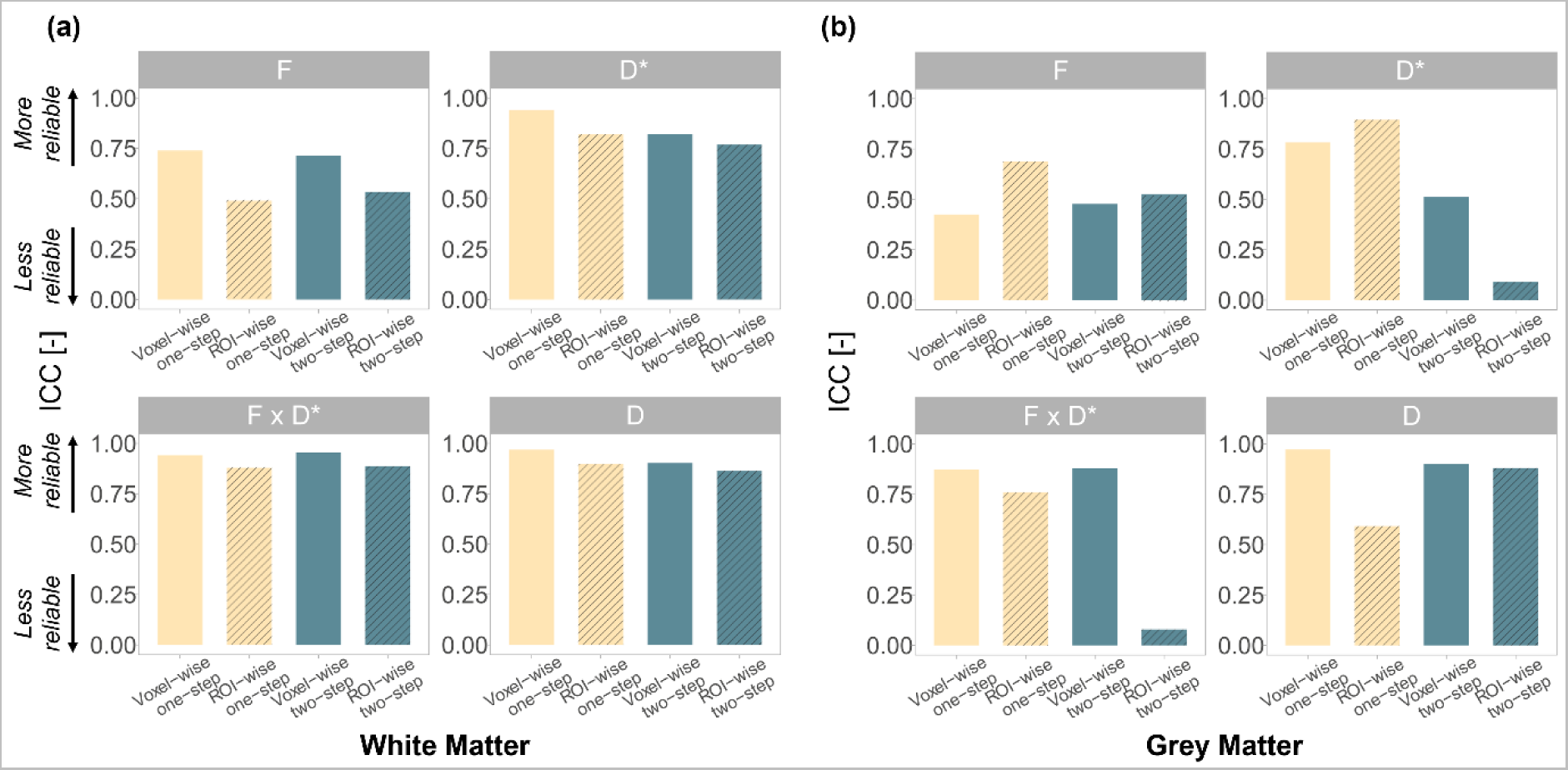
ICC values of the IVIM parameters obtained using the voxel-wise and ROI-wise fits and both the one-step and two-step algorithms in the (a) white matter and (b) grey matter.

The Bland-Altman analysis yielded bias and limits of agreement, which are reported for both voxel-wise and ROI-wise fits using the one-step and two-step algorithms in Supplementary Table S1. The Bland-Altman plots revealed narrower limits of agreement using the one-step method compared to the two-step when fitting the signal in a voxel-wise manner, except for *F* · *D\**, in the white and grey matter (Figure 5).

**Figure 5:**
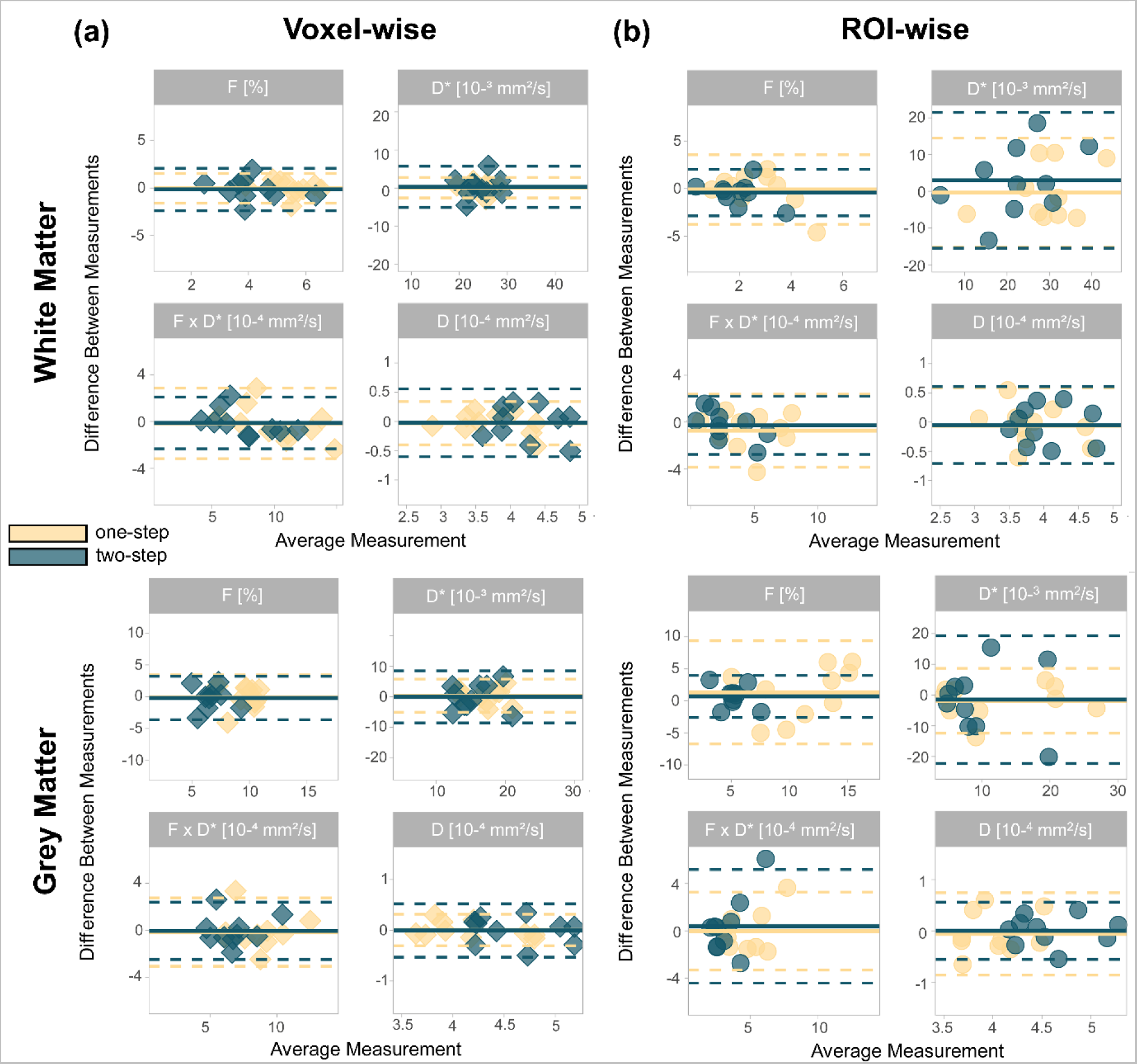
Bland-Altman plots of the IVIM parameters in the white matter (first row) and grey matter (second row) obtained using the (a) the voxel-wise and (b) ROI-wise fit with the one-step and two-step algorithms. The bold lines represent the mean difference between the two sessions (bias) and dashed lines represent the 95% limits of agreement of the mean difference.

IVIM parameters showed good correlations between first and second scan using the voxel-wise one-step fitting approach for *D\**, *F* · *D\** and *D* in the white matter (Pearson’s correlation coefficient ≥ 0.88) (Figure 6). The correlations were overall stronger using the voxel-wise fit compared to ROI-wise, and with the one-step approach compared to the two-step one. In general, *F* showed a more moderate inter-session correlation with all fitting approaches (WM: 0.35 ≤ R ≤ 0.57, GM: 0.25 ≤ R ≤ 0.54).

**Figure 6:**
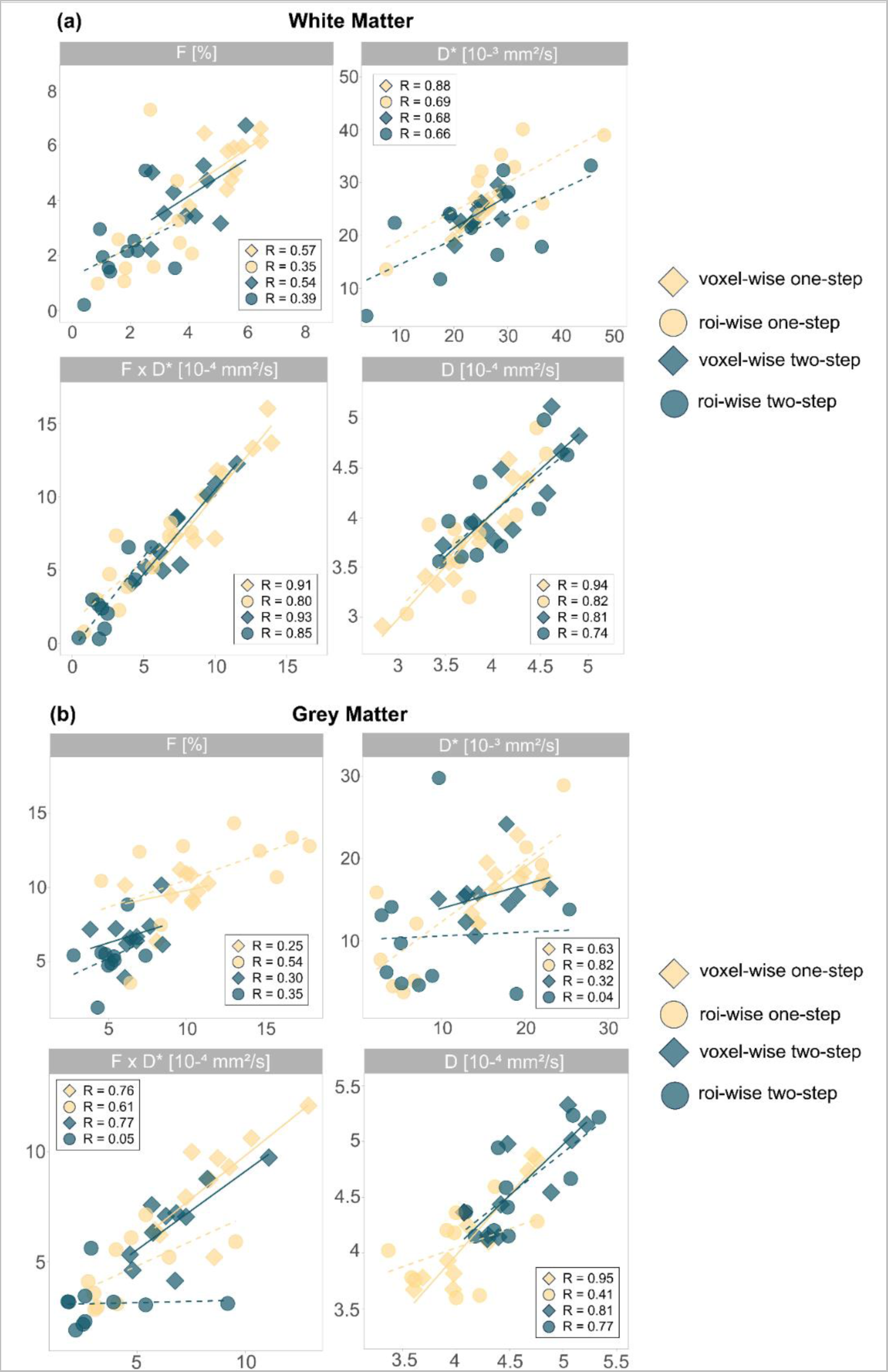
Scatterplots of the IVIM parameters and linear correlations between the first and second sessions in the (a) white matter and (b) grey matter with the voxel-wise fit (bold line) and the ROI-wise fit (dashed line), using the one-step and two-step algorithms, along with the Pearson correlation coefficient R.

## Discussion

This study showed high repeatability and reliability of IVIM parameters calculated in the cervical cord applying different fitting approaches. The voxel-wise fitting approach demonstrated higher within-subject repeatability (i.e., lower CVs and higher ICC values) for the IVIM parameters compared to the ROI-wise method. While the one-step tended to improve the repeatability of IVIM parameters compared with the two-step method, the difference was not found significantly different.

### IVIM parameters in the cervical cord and repeatability

Microvascular volume fraction (*F*) maps visually showed higher intensity in the anterior and intermediate grey matter sub-regions of the cord, which is in line with microangiography data of the cord vasculature^44^ and previous studies^11,45^. This study showed a good-to-excellent reliability of IVIM parameters *D\**, *F* · *D\**, and *D*, in the cervical cord using a voxel-wise (one-step) fit.

### Comparing voxel-wise and ROI-wise fitting approaches

The tissue diffusion coefficient (*D*) remained similar in both the voxel-wise and ROI-wise fitting approaches, attesting of the robustness of the estimation of *D* to the choice of the fitting method^46^. On the other hand, *F* and *D\** values differed depending on the fitting approach applied, with the ROI-wise method resulting in lower *F* (range of average values with voxel-wise: 4.03 – 5.49, ROI-wise: 1.73 – 2.76) and more variation in *D\** (range of average values with voxel-wise: 24.07 – 24.41, ROI-wise: 21.23 – 29.57), including one-step and two-step, in the white matter. This finding suggests a higher variability in the signal attenuation at lower b-values and demonstrates the importance of the fit choice to model the perfusion contribution to the signal.

The voxel-wise fit showed higher repeatability performance compared to the ROI-wise fit, with significantly lower within-subject CVs (WM: *F*: p = 0.001, *D\**: p = 5.3e-5, *F* · *D\**: p = 0.005; GM: *D\**: 0.0003, *F* · *D\**: p = 0.002, *D*: p = 0.01) and higher ICC values, indicating the sensitivity of the fit quality on the fitting approach. The higher variability of the ROI-wise fitting approach may be due to varying IVIM values within the region where the signal is averaged across. Additionally, partial volume effects between grey matter, white matter, and CSF regions, as well as between different vessel sizes (capillaries to arteries) may lead to more variability. The theoretical advantage of the ROI-wise fitting approach is the mitigation of the noise at the input of the fitting algorithm. However, given the large number of repetitions used for each b-value (20/b-value), the SNR is unlikely to be a limitation for the voxel-wise fitting approach, as observed with the high range of SNR values obtained, which were of a similar range of SNR values obtained at 7T in a previous study^11^. Of note, the currently lower repeatability of the ROI-wise fitting compared to the voxel-wise fit may change for acquisitions with a lower SNR (e.g., with less repetitions per b-value). Using the voxel-wise fit, the white matter showed better repeatability measures compared to the grey matter. This is also supported by lower average CVs, higher ICC values, and smaller mean bias with narrower limits of agreements in the Bland-Altman analysis. The results are in line with the strong linear correlation coefficients between the IVIM values in the first and second session. The overall lower repeatability observed in the grey matter is most certainly due to the influence of partial volume effect, small number of voxels, and possible registration imprecisions occurring during the processing steps.

### Comparing one-step and two-step fitting algorithms

The differences between the one-step and two-step fitting approaches were not found significant. However, previous reports suggested the segmented method (e.g., two-step) to be beneficial at lower SNR for *D*, whereas the one-step method may yield better reliability in cases of higher SNR^47,48^. Those results were also observed in our study, where the one-step fitting approach showed higher reliability (higher ICC) compared to the two-step with the ROI-wise fit, in which the effective SNR is higher due to the average of signal across the region. The difference between the one-step and two-step methods applying the voxel-wise fitting approach was not significant, yet the one-step led to higher ICC in the white matter compared to the two-step. Individual CV values up to 30% were observed for the voxel-wise one-step fitting method, suggesting that, although the results are promising at the group level, there is room for improvement in terms of confidence at the individual level, which should be kept in mind during the interpretation of clinical findings.

Of note, a clinical study has investigated perfusion impairment in patients with degenerative cervical myelopathy compared to healthy subjects, using the current IVIM protocol^49^. The results showed a significant decrease in *D\** and *F* · *D\**, with an average difference of -9.5% and -15.9% in the white matter and -11.0 % and -14.4% in the grey matter, respectively, at the group level (29 DCM patients, 30 healthy subjects), using the voxel-wise one-step fit. In comparison, the average test-retest differences observed for the parameters in the current study (considering the whole group) were in the range between 4.39% and 11.66% in the white matter, and 12.77% and 12.12% in the grey matter, while the maximum test-retest differences observed at the individual level were 12.15%, 17.37% and 26.67%, 32.88%, respectively. Moreover, the differences observed in patients give confidence that the estimation method does not compress or restrain the true variations of the parameters while ensuring a good test-retest repeatability.

Furthermore, our results in the cervical cord show high repeatability compared to previous studies on IVIM reliability which used different acquisition protocols and fitting approaches in different organs^46,50–53^. A previous study applying IVIM in head and neck tissues reported CVs of *F* and *D\** varying between 15.27% and 22.14 %, and 29.24% and 41.80%, respectively^51^. In lung cancer, the mean CVs of *F* ranged between 36.54% and 38.34%, and 68.59% and 72.62% for *D\**, while *D* had lower CV (11-12% - 11.33%)^50^. Across previous studies, *D* parameter overall showed a better repeatability, which is in line with our results.

### Comparison to different fitting methods

In the recent years, extensive effort has been made in investigating alternative approaches for the fitting of IVIM data, such as Bayesian methods^7^ and deep learning-based algorithms^52^, which have shown great promises in terms of noise robustness and parameter estimation accuracy. The study of those alternative methods was beyond the scope of the current study and would necessitate further technical development.

### Limitations

This study has a few limitations. First, the sample size and the number of sample points were rather small, and larger cohorts and more retests are required to provide more accuracy. However, the current cohort size is comparable to previous IVIM reliability studies^51–54^. Second, young healthy volunteers (24 years < age < 42 years) were recruited in this study. Repeatability with patients and elderly cohorts can be lower, which should be either accounted for in clinical studies or quantified in subsequent research. Furthermore, the nominal acquisition time of the protocol is currently long (ca. 39 minutes) to be transferred to clinical routine. The acquired test-retest IVIM maps are a great opportunity to perform time optimization of the acquisition protocol without compromising the obtained SNR nor the reliability of the IVIM values. It must also be noted that the test-retest variability reported here includes the test-retest variability of the entire post-processing chain, including the automatic segmentation and calculation of the probabilistic masks. The different steps involved are not free from inter-session variations and might contribute to the final test-retest variability of the IVIM values reported in this study.

## Conclusion

This study showed high repeatability of IVIM parameters in the human cervical cord assessed with four different fitting approaches, comparing voxel-wise vs. ROI-wise fits, and one-step vs. two-step fitting algorithms. The voxel-wise fitting approach showed a higher repeatability of IVIM-derived parameters in the cervical cord, compared to the ROI-wise fit approach, independently of using the one-step or two-step algorithms. The present work provides hereby reference values for future clinical investigations of the perfusion integrity in the cervical cord in different neurodegenerative pathologies.

## Data and Code Availability

Anonymized datasets used and analyzed during the current study are available from the corresponding author on reasonable request.

## Acknowledgements

The authors would like to thank all the participants who volunteered to take part in this study. This work is based on experiments performed at the Swiss Center for Musculoskeletal Imaging (SCMI), Balgrist Campus AG, Zurich, and methodological developments performed at the Centre de Résonance Magnétique Biologique et Médicale, CRMBM-CEMEREM, Aix-Marseille University/CNRS, Marseille, France.

